# A Simple, Home-Therapy Algorithm to Prevent Hospitalization for COVID-19 Patients: *A Retrospective Observational Matched-Cohort Study*

**DOI:** 10.1101/2021.03.25.21254296

**Authors:** Fredy Suter, Elena Consolaro, Stefania Pedroni, Chiara Moroni, Elena Pastò, Maria Vittoria Paganini, Grazia Pravettoni, Umberto Cantarelli, Nadia Rubis, Norberto Perico, Annalisa Perna, Tobia Peracchi, Piero Ruggenenti, Giuseppe Remuzzi

**Author notes:** Correspondence to:* Norberto Perico MD, Istituto di Ricerche Farmacologiche IRCCS, Centro di Ricerche Cliniche per Malattie Rare Aldo e Cele Daccò, Via GB Camozzi 3, 24020 Ranica, Bergamo, Italy. equally contributed (Norberto Perico, Piero Ruggenenti, Giuseppe Remuzzi).

## Abstract

**Background:** Effective home treatment algorithms implemented based on a pathophysiologic and pharmacologic rationale to accelerate recovery and prevent hospitalisation of patients with early coronavirus disease 2019 (COVID-19) would have major implications for patients and health system.

**Methods:** This academic, matched-cohort study compared outcomes of 90 consecutive consenting patients with mild COVID-19 treated at home by their family physicians between October 2020 and January 2021, according to the proposed recommendation algorithm, with outcomes for 90 age-, sex-, and comorbidities-matched patients who received other therapeutic regimens. Primary outcome was time to resolution of major symptoms. Secondary outcomes included prevention of hospitalisation. Analyses were by intention-to-treat.

**Findings:** All patients achieved complete remission. The median [IQR] time to resolution of major symptoms was 18 [14-23] days in the ‘recommended’ schedule cohort and 14 [7-30] days in the matched ‘control’ cohort (p=0·033). Other symptoms persisted in a lower percentage of patients in the ‘recommended’ than in the ‘control’ cohort (23·3% versus 73·3%, respectively, p<0·0001) and for a shorter period (p=0·0107). Two patients in the ‘recommended’ cohort were hospitalised compared to 13 (14·4%) controls (Log-rank test, p=0·0038). The prevention algorithm reduced the days and cumulative costs of hospitalisation by >90% (from 481 to 44 days and from €296.000 to €28.000, respectively. 1.2 patients had to be treated to prevent one hospitalisation event.

**Interpretation:** Implementation of an early home treatment algorithm failed to accelerate recovery from major symptoms of COVID 19, but almost eliminated the risk of hospitalisation and related treatment costs.

**Research in Context:** *Evidence before this study:* We searched PubMed and the Cochrane Library for peer-reviewed articles published in any language up to March 19, 2021, using the search terms “2019-nCoV” or “SARS-CoV-2” or “COVID-19” and “early” or “outpatient” or “treatment” or “home”. Our search did not identify any randomised clinical trials or observational studies that assessed the effectiveness of treatment regimens targeting early, mild symptoms of COVID-19 in the outpatient setting.

*Added value of this study:* In this fully academic, observational matched-cohort study, we found that early home treatment of 90 consecutive patients with mild COVID-19 by their family physicians according to the proposed recommendation algorithm, designed based on a pathophysiologic and pharmacologic rationale, required few more days to achieve resolution of major symptoms including fever, dyspnea, musculoskeletal pain, headache and cough compared to 90 age-, sex-, and comorbidities-matched patients who received other therapeutic regimens (primary outcome). Nonetheless, it is noteworthy that the home treatment of COVID-19 patients according to the proposed recommendation algorithm significantly reduced the risk of hospitalization compared to the other treatments in the ‘control’ cohort. Days of hospitalization and related treatment costs were reduced by over 90% in the ‘recommended’ cohort as compared to ‘control’ cohort. Just 1.2 patients needed to be treated according to the recommendation algorithm to prevent one hospitalization event. We also found that symptoms such as anosmia and ageusia/dysgeusia were less persistent and lasted a shorter time in the ‘recommendation’ than in the ‘control’ cohort.

*Implications of the available evidence:* The finding that the implementation of the proposed simple treatment algorithm during the initial, mild phase of COVID-19 has the potential to prevent disease progression, potentially limiting the need for hospital admission, may have major implications for patients and health care providers. Indeed, preventing hospitalisations due to the worsening of COVID-19 will not only save lives, but will also contribute to remarkably reduced treatment costs and to streamlining health care systems that are overburdened by the effects of the pandemic. However, time to hospitalization was a secondary outcome of the study and the possibility of a casual finding cannot be definitely excluded. Thus, the observed reduction in patients hospitalizations should be considered as an hypothesis generating finding that could provide a robust background for a prospective trial primarily aimed to test treatment effect on this outcome.

## Introduction

The newly recognised disease COVID-19 is caused by the Severe-Acute-Respiratory-Syndrome Coronavirus 2 (SARS-CoV-2), which rapidly spread globally in late 2019, reaching pandemic proportions.^1^ The clinical spectrum of SARS-CoV-2 infection is broad, encompassing asymptomatic infection, mild upper respiratory tract illness and mild extrapulmonary symptoms, and severe viral pneumonia with respiratory failure and even death.^2,3^ Given the rising global death toll associated with the pandemic,^1^ in the last year we have witnessed a race to find drugs/biological treatments to save the lives of hospitalised, severely ill patients, as well as to develop vaccines.^4,5^ To this end, randomised clinical trials have been performed or are underway to test experimental drug candidates and repurposed medicines.^6,7^ Nonetheless, to limit the number of hospitalisations and deaths due to severe illness, thus avoiding pushing hospitals to their limits and remarkably reducing the tremendous treatment costs for health care providers,^8^ it is crucial to also focus on primary care physicians and initial mild symptoms in COVID-19 patients at home.

As with other acute viral infections, early initiation of treatment for COVID-19 might improve clinical outcomes.^9^ For COVID-19, most primary care physicians have initially treated their patients according to their judgement, with various treatment regimens they believe are most appropriate based on their experience/expertise. We recently published a note on how we were treating patients at home based on the pathophysiologic and pharmacologic rationale and the available clinical evidence of efficacy in COVID-19 for each of the recommended class of drugs.^10^ This consists of anti-inflammatory agents, especially relatively selective cyclooxygenase-2 (COX-2) inhibitors,^11^ given early in the course of the disease at the very beginning of the onset of symptoms, even before the nasopharyngeal swab, an approach that is intended to limit excessive host inflammatory responses to viral infection.^10^ Others have debated the same issue for corticosteroids ^12^ and also mentioned the risk of secondary infections and other complications.

Moreover, COVID-19 patients are exposed to the risk of thromboembolic events, and anticoagulant prophylaxis is recommended, unless contraindicated.^13,14^ However, no randomised clinical trials have been performed so far in COVID-19 patients to compare the effectiveness of different regimens targeting early symptoms at home. Comparative analysis of patient cohorts in everyday clinical practice with adjustment for possible confounding bias may offer a good alternative to randomised clinical trials to evaluate the effectiveness of novel therapies.^15,16^ Thus, we used this approach in a retrospective observational matched-cohort study to compare the outcomes of a cohort of COVID-19 patients treated at home by their family physicians according to a therapeutic paradigm based on the proposed recommendations ^10^ with the outcomes of a cohort of similar patients treated with other therapeutic regimens.

## Patients and Methods

### Study design and participants

This retrospective observational study included two matched cohorts of COVID-19 patients. The ‘recommended schedule’ cohort included 90 patients treated at home by their family doctors according to published proposed recommendations ^10,17^ between October 2020 and January 2021. It involved family physicians from the Bergamo, Varese and Teramo provinces who had followed the proposed recommendations and expressed their interest in participating in the study with the engagement of their patients. They applied the recommended treatment algorithm (see Supplementary Methods) at the onset of, or within a few days of, the onset of symptoms. The doctors were asked to complete an online questionnaire after collecting the consent form signed by the patients. To this end, patients received detailed information from their physicians on the objectives and design of the study. The questionnaire included information on the outcomes of COVID-19 symptoms/illness that were relevant to addressing the primary, secondary and safety aims of the observational study. The study coordinator, the Istituto di Ricerche Farmacologiche Mario Negri IRCCS, promoted the project through online institutional media. Male and female adults (≥ 18 years old), with early, mild symptoms of COVID-19, who started the recommended treatment without waiting for the results of a nasopharyngeal swab, if any, were eligible to participate. Subjects who required immediate hospital admission because of severe COVID-19 symptoms at onset, according to the family doctor’s assessment, were excluded.

Ninety COVID-19 patients matched by age, sex, concomitant diseases (hypertension, diabetes, cardiovascular diseases, obesity, chronic kidney disease) and symptoms at onset of illness, who had been enrolled in the “Study of the Genetic Factors That Influence the Susceptibility to and Severity of COVID-19” (the ORIGIN study) and treated at home by family physicians with drug regimens that were not necessarily guided by those proposed in the recommendations, served as controls. In this cohort, too, individuals who needed immediate hospitalisation according to the family physician’s assessment because of severe symptoms of illness at onset, were not included. ORIGIN is a large study being conducted by the Istituto di Ricerche Farmacologiche Mario Negri IRCCS with the general aim of exploring whether variations in inter-individual genetic signature in the population of COVID-19 patients living in the Bergamo province could explain the observed different responses to SARS-CoV-2 viral infection and thus different clinical features of the disease (ClinicalTrials.gov; NCT04799834). ORIGIN collects, among other types of information, all clinical information planned for the analysis of the ‘recommended schedule’ cohort. So far over 5000 consenting subjects have joined the ORIGIN study.

The COVER study has been approved by the Centralised Ethical Committee for all COVID-19 trials in Italy based at the Istituto Nazionale per le Malattie Infettive Lazzaro Spallanzani, Rome (Parere n° 263, January 31, 2021) and registered at the ClinicalTrials.gov (NCT04794998). All COVER participants provided written informed consent to participating in the study.

### Outcomes and definitions

The primary outcome was the time (in days) from beginning the proposed recommended treatments or other therapeutic regimens to resolution of major symptoms (time to complete remission). “Complete remission” was defined as complete recovery from major symptoms, i.e. no fever, dyspnoea and/or SpO2 >94%, cough, rhinitis, pain (myalgia, arthralgia, chest pain, headache, sore throat), vertigo, nausea, vomiting or diarrhoea, nor sicca syndrome or red eyes.

Secondary outcomes included: 1) Days between the onset of symptoms and the start of anti-inflammatory therapy in the two treatment cohorts. 2) Compliance with the algorithm in the cohort adopting the proposed treatment recommendations, defined as adherence to recommended schedule, daily dose of drugs and duration of treatment. 3) Rate of complete remission, as defined above, in the two treatment cohorts. 4) Rate of remission with persistence of very mild symptoms in the two cohorts. This was termed “partial remission”, and defined as recovery from major COVID-19 symptoms, but persistence of symptoms such as anosmia, ageusia/dysgeusia, lack of appetite, fatigue. In addition, time of persistence of these symptoms (<30 days, or 30 to 60, or >60 days after “complete remission”) was assessed. 5) Rate of patients worsening with severe dyspnoea requiring hospitalisation in the two treatment cohorts.

We predefined potential baseline confounders, such as age, sex, and concomitant diseases that potentially enhance the risk of severe COVID-19 illness.^18–20^

In addition, serious (SAE) and non-serious adverse events (AE) related to the administered treatments were assessed. The severity/non-severity of the observed events and their causal relationships with treatments were determined by the family doctor in charge of the patients.

### Sample size and statistical analyses

Given the results of a recently published study ^21^ and considering the characteristics of our COVID-19 patient population, we assumed that our ‘control cohort’ may have a longer time to resolution of symptoms (time to complete remission), expected to be equal to 20 days (SD: 10 days) and that in the ‘recommended schedule’ cohort it would be shortened to 15 days. With the above assumptions, a sample size of 86 per group (172 total) would achieve 90% power to reject the null hypothesis of equal means when the population mean difference is μ1 - μ2 = 20 - 15 = 5 days with a standard deviation for both groups of 10 days and with a significance level (alpha) of 0.05 using a two-sided two-sample equal-variance t-test. Accounting for a 20% drop-out rate, 108 per group (i.e. 216 total) needed to be included.

‘Recommended schedule’ and ‘control’ cohorts were expected to be sufficiently comparable at baseline. However, matching was carried out between the two groups. ^22^ Scores were built with logistic regression by using the “Propensity Score” SAS procedure, which considered at least the following baseline variables: age, sex, comorbidities, and COVID-19 symptoms at onset. Continuous variables were analysed through descriptive statistics and reported as mean (SD) or median [IQR], as appropriate. Within-group changes with respect to baseline were analysed with paired t-test or Wilcoxon signed-rank test, as appropriate. For survival data a Log-rank test was used. For the primary outcome a p-value of 0.05 was considered to determine statistical significance. For the five secondary outcomes a Bonferroni-adjusted p-value of 0.01 was used.

## Results

Between October 2020 and January 2021, seven family physicians who expressed an interest in participating in this retrospective study reported 90 consecutive consenting participants with early symptoms of COVID-19 whom they treated at home according to the proposed recommendations (‘recommendation’ cohort).^10^ All these individuals had confirmed SARS-CoV-2 infection by positive nasopharyngeal swabs. Eighty-eight of the 90 individuals identified from the ORIGIN dataset who had been matched for age, sex, and major concomitant diseases (‘control’ cohort) presented with COVID 19 between March and May 2020 and two participants in October 2020 and January 2021. All were COVID-19 cases confirmed by nasopharyngeal swab or by serology tests, and treated at home by their family doctors with whatever regimen the doctor believed was most appropriate based on their expertise/experience. Both cohorts had a slight prevalence of females (56·7%) and were comparable in terms of age range, with most individuals aged between 41 and 65 (Table 1). Similarly, the distribution of concomitant diseases was well balanced between the two groups, with a few more individuals with hypertension and chronic kidney disease in the ‘recommended’ cohort than in the ‘control’ cohort. The most common symptoms at the onset of disease were musculoskeletal pain (91·1% vs 83·3%) and fever (80·0% vs 78·9%), followed by fatigue (73·3% vs 76·7%), cough (60·0% vs 45·6%) and headache (56·7% vs 41·1%) in both cohorts (Table 1). More patients in the ‘recommended’ cohort had rhinitis at onset (26·7% vs 8·9%, p=0.003), while diarrhoea (14·4% vs 30·0%, p=0·019) and dyspnoea (20·0% vs 36·7%, p=0·02) were significantly more frequent in the ‘control’ cohort. On average, dyspnoea occurred 4 to 5 days after the onset of symptoms in the ‘recommended’ cohort.

**Table 1.** Demographic and early symptoms associated with COVID-19 illness in the two treatment cohorts.

|  | **Overall**<br>(*n*=180) | **Recommended**<br>**treatment**<br>**cohort**<br>(*n*=90) | **Control**<br>**cohort**<br>(*n*=90) | **P value** |
| --- | --- | --- | --- | --- |
| **Demographic characteristics** |  |  |  |  |
| Age, years |  |  |  |  |
| 18-40 | 34 (18.89) | 17 (18.89) | 17 (18.89) | 1.000 |
| 41-65 | 90 (50.00) | 45 (50.00) | 45 (50.00) |  |
| 66-75 | 26 (14.44) | 13 (14.44) | 13 (14.44) |  |
| >75 | 30 (16.67) | 15 (16.67) | 15 (16.67) |  |
| Males, *n* (%) | 78 (43.33) | 39 (43.33) | 39 (43.33) | 1.000 |
| **Comorbidities, *n* (%)** |  |  |  |  |
| Cardiovascular disease | 32 (17.78) | 16 (17.78) | 16 (17.78) | 1.000 |
| Hypertension | 57 (31.67) | 31 (34.44) | 26 (28.89) | 0.522 |
| Diabetes mellitus | 16 (8.89) | 8 (8.89) | 8 (8.89) | 1.000 |
| Overweight/Obesity | 31 (17.22) | 16 (17.78) | 15 (16.67) | 1.000 |
| Chronic kidney disease | 2 (1.11) | 2 (2.22) | 0 (0) | 0.497 |
| **Early symptoms, *n* (%)** |  |  |  |  |
| Fever | 143 (79.44) | 72 (80.00) | 71 (78.89) | 1.000 |
| Myalgia | 100 (55.56) | 50 (55.56) | 50 (55.56) | 1.000 |
| Arthralgia | 57 (31.67) | 32 (35.56) | 25 (27.78) | 0.336 |
| Tiredness/exhaustion | 135 (75.00) | 66 (73.33) | 69 (76.67) | 0.731 |
| Dyspnea | 51 (28.33) | 18 (20.00) | 33 (36.67) | 0.020 |
| Chest pain | 23 (12.78) | 10 (11.11) | 13 (14.44) | 0.656 |
| Headache | 88 (48.89) | 51 (56.67) | 37 (41.11) | 0.052 |
| Lack of appetite | 68 (37.78) | 28 (31.11) | 40 (44.44) | 0.090 |
| Cough | 95 (52.78) | 54 (60.00) | 41 (45.56) | 0.073 |
| Sore throat | 37 (20.56) | 22 (24.44) | 15 (16.67) | 0.268 |
| Rhinitis | 32 (17.78) | 24 (26.67) | 8 (8.89) | 0.003 |
| Vomiting/nausea | 34 (18.89) | 13 (14.44) | 21 (23.33) | 0.182 |
| Diarrhoea | 40 (22.22) | 13 (14.44) | 27 (30.00) | 0.019 |
| Red eyes | 20 (11.11) | 7 (7.78) | 13 (14.44) | 0.235 |
| Vertigo | 5 (2.78) | 3 (3.33) | 2 (2.22) | 1.000 |
| Sicca syndrome | 3 (1.67) | 0 (0) | 3 (3.33) | 0.246 |
| Anosmia | 100 (55.56) | 46 (51.11) | 54 (60.00) | 0.294 |
| Ageusia | 102 (56.67) | 45 (50.00) | 57 (63.33) | 0.098 |

Data are numbers (percentages). Between-group differences were assessed by Fisher's exact test.

### Primary outcome

The median time to resolution of major symptoms (complete remission) was 18 days [IQR: 14-23] in the ‘recommendation’ cohort, slightly but significantly longer (p=0·033) than in the matched ‘control’ cohort (14 days, IQR: 7-30) (Figure 1 A). Time to complete remission was comparable between females (median [IQR], ‘recommended’ cohort: 18 days [14-23]; ‘control’ cohort: 15 days [8-30], p=0·116) and males (median [IQR], ‘recommended’ cohort: 16 days [12-23]; ‘control’ cohort: 10 days [6-30], p=0·128) of the two cohorts (Figure 1B). Similarly, there was no significant difference regarding time to complete remission between the two cohorts for patients under the age of 65. The median time to resolution was, however, significantly longer in the ‘recommended’ than in the ‘control’ cohort for elderly individuals (> 66 years old) (Figure 1C).

**Figure 1.**
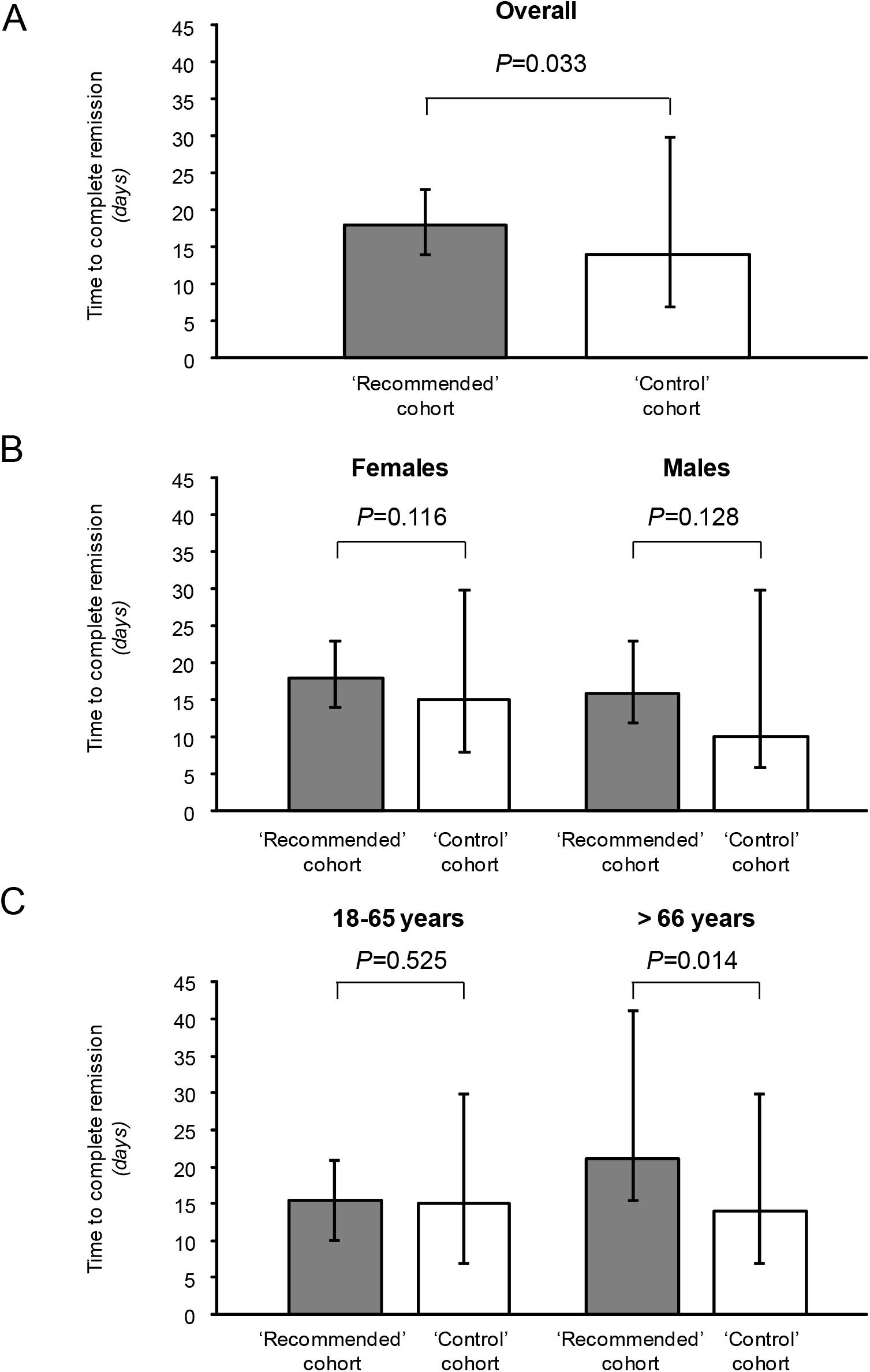
Time to complete remission. Time to complete remission in the two treatment cohorts (primary outcome, Panel A), in the two treatment cohorts according to sex (Panel B), and in the two treatment cohorts according to age range (Panel C). Data are median and interquartile range. Grey histograms, recommended treatment cohort; white histograms, control cohort. Between-group differences were assessed by Mann-Whitney test.

### Secondary outcomes

Two of the 90 patients (2·2%) in the ‘recommended’ cohort were hospitalised, compared to 13 of the 90 (14·4%) in the ‘control’ cohort (Figure 2A). In the ‘recommended’ cohort one patient was hospitalised due to interstitial pneumonia (Table 2). However, he spontaneously started taking paracetamol at home before contacting his doctor, which must be considered a protocol violation. The other patients in this cohort were admitted to hospital 11 days after complete remission of COVID-19 symptoms and SARS-CoV-2 negative nasopharyngeal swab, due to dyspnoea developed a few days after right frontal lobe trauma during a post syncopal episode that was related to a documented pulmonary embolism (Table 2). All patients in the ‘control’ cohort were hospitalised due to dyspnoea secondary to interstitial pneumonia (Table 2). The event rate was significantly lower in the ‘recommended’ than in the ‘control’ cohort (Log-rank test, p=0·0038) (Figure 2A). The median [IQR] of days of hospitalisation was numerically lower in the ‘recommended’ than in the ‘control’ cohort (22.0 days [7.0-37.0] vs 32.5 days [15-0-56.5], p=0.465) (Table 2). The cumulative number of days in the ICU, in sub-intensive care units, and ordinary units were, respectively, 11, 1, and 32 in the ‘recommended’ cohort, and 104, 13, and 364 in the ‘control’ cohort (Figure 3A). Thus, overall, there were only 44 days of hospitalisation in the “recommended” cohort, compared to 481 in controls (9.1%). Consistently, cumulative hospitalisation costs were €28,335 vs €296,243 for controls (9.6%) (Figure 3B). Only 1.2 [95% CI: 1.1 to 1.3] patients needed to be treated with the home therapy algorithm to prevent one hospitalisation event.

**Table 2.** Clinical course of hospitalised patients in the two cohorts.

| Cohort | Reason for hospital admission | Hospitalisation (days) | Oxygen therapy* (yes/no) | CPAP (yes/no) | CPAP (days) | Mechanical ventilation (yes/no) | Mechanical ventilation (days) | ICU admission (yes/no) | ICU admission (days) | Sequelae at discharge (yes/no) |
| --- | --- | --- | --- | --- | --- | --- | --- | --- | --- | --- |
| *Control* |  |  |  |  |  |  |  |  |  |  |
| Control | Dyspnea (interstitial pneumonia) | 60 | Yes | Yes | 3 | Yes | 17 | Yes | 17 | No |
| Control | Dyspnea (interstitial pneumonia) | 8 | Yes | No | - | No | - | No | - | No |
| Control | Dyspnea (interstitial pneumonia) | 5 | Yes | No | - | No | - | No | - | No |
| Control | Dyspnea (interstitial pneumonia) | 68 | Yes | Yes | 3 | Yes | 14 | Yes | 14 | Yes, persistence of decreased muscle tone |
| Control | Dyspnea (interstitial pneumonia) | 10 | Yes | No | - | No | - | No | - | No |
| Control | Dyspnea (interstitial pneumonia) | 41 | Yes | No | - | Yes | 25 | Yes | 25 | No |
| Control | Dyspnea (interstitial pneumonia) | 35 | Yes | No | - | No | - | No | - | No |
| Control | Dyspnea (interstitial pneumonia) | 23 | Yes | No | - | No | - | No | - | No |
| Control | Dyspnea (interstitial pneumonia) | 50 | Yes | No | - | No | - | No | - | No |
| Control | Dyspnea (interstitial pneumonia) | 20 | Yes | No | - | No | - | No | - | No |
| Control | Dyspnea (interstitial pneumonia, small pulmonary embolism in stila bronches of the right lung) | 128 | Yes | Yes | 7 | Yes | 48 | Yes | 48 | Yes, gait disturbances in polyneuropathy, severe chronic respiratory insufficiency |
| Control | Dyspnea (interstitial pneumonia) | 30 | Yes | No | - | No | - | No | - | No |
| Control<sup>o</sup> | Dyspnea (interstitial pneumonia) |  | Yes | No | - | No | - | No | - |  |
| *'Recommended'* |  |  |  |  |  |  |  |  |  |  |
| 'Recommended' | Dyspnea (massive bilateral pulmonary embolism and left iliac-femoral deep vein thrombosis, after right frontal lobe trauma post-syncopal episode) | 7 | Yes | No | - | No | - | No | - | Yes, persistence of pulmonary thromboembolism, venous thrombosis in resolution. |
| 'Recommended' | Dyspnea (interstitial pneumonia) | 37 | Yes | Yes | 1 | Yes | 11 | Yes | 11 | No |

\*Conventional oxygen therapy (oxygen delivered by nasal tube, nasal cannula or face mask). <sup>o</sup> This patients did not provide the hospital discharge letter. CPAP, continuous positive airway pressure; ICU, intensive care unit.

**Figure 2.**
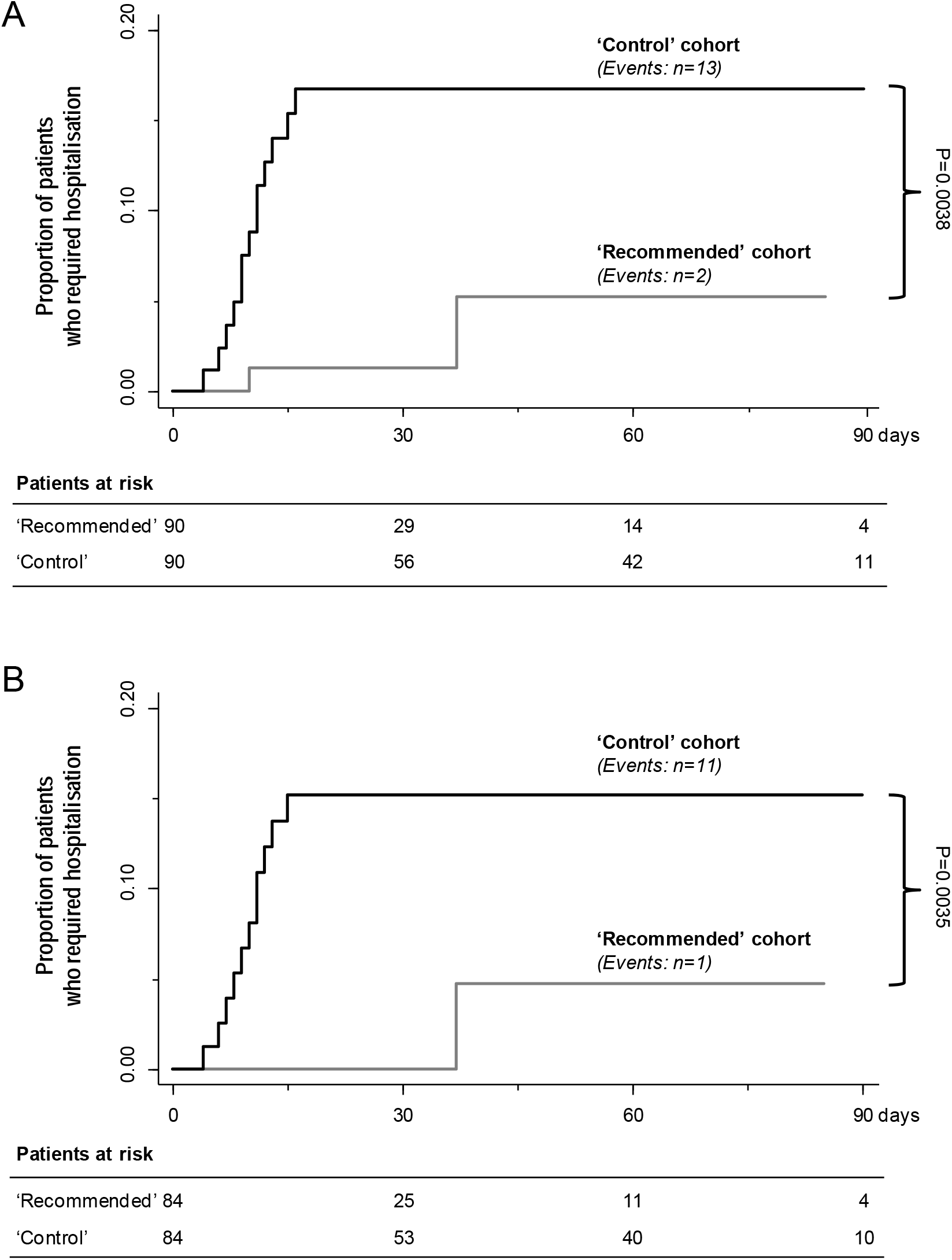
Kaplan-Meier curves for hospital admission. Kaplan-Meier curves show the proportion of patients who required hospitalisation in the two treatment cohorts (Panel A), and after excluding patients who spontaneously started treatment with paracetamol before contacting their family doctors in the “recommended” cohort and the related matched patients in the “control” cohort (Panel B). Grey line, recommended treatment cohort; black line, control cohort. Between-group differences were assessed by Log-rank test.

**Figure 3.**
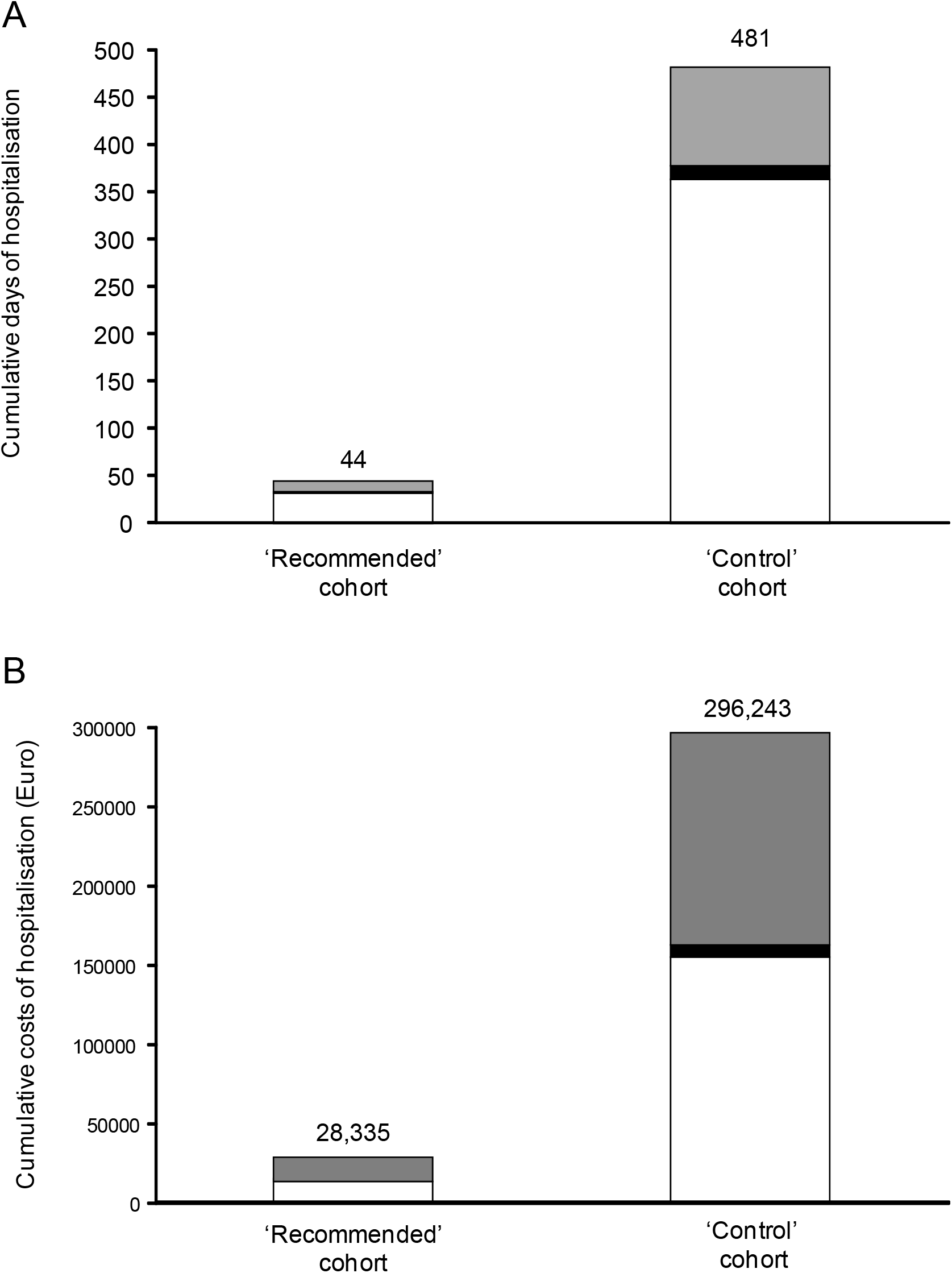
Cumulative days of hospitalisation and related costs in the two study cohorts. Cumulative days of hospitalisation in the ‘recommended’ treatment cohort and in the ‘control’ cohort, according to stay in ordinary ward (white), subintensive care unit (black) and intensive care unit (grey) (Panel A). Cumulative costs for hospitalisation in the ‘recommended’ treatment cohort and in the ‘control’ cohort, according to stay in ordinary ward (white), subintensive care unit (black) and intensive care unit (grey) (Panel B).

In the ‘recommended’ cohort, 66 of 90 patients were given a relatively selective COX-2 inhibitor (nimesulide or celecoxib) (Table 3). Twenty patients received other NSAIDs, including aspirin (n=7). Thirteen patients were prescribed ibuprofen or indomethacin or acetaminophen (paracetamol), bringing non-adherence to the recommended anti-inflammatory regimen to 14·4% in the cohort (Table 3). On the other hand, in the ‘control’ cohort, none of the patients received relatively selective COX-2 inhibitors and only one was given aspirin (Table 3). Moreover, in this cohort, most patients were treated with paracetamol (n=45), and the remaining with ketoprofen or ibuprofen. Thirty percent of patients in the ‘recommended’ cohort and 9·2% in the ‘control’ cohort were given corticosteroids (p=0·001) (Table 3). More patients were prescribed antibiotics (p<0·001) as well as anticoagulants (p=0·004) in the ‘recommended’ than in the ‘control’ cohort (Table 2). Regarding antibiotic therapy, in the ‘recommended’ cohort, 49% of treated patients were given azithromycin and 15·7% amoxicillin/clavulanic acid. Seven patients in the ‘recommended’ cohort and six in the ‘control’ cohort required gentle oxygen supply at home for decreasing oxygen saturation or following a first episode of dyspnoea or wheezing (Table 3).

**Table 3.**
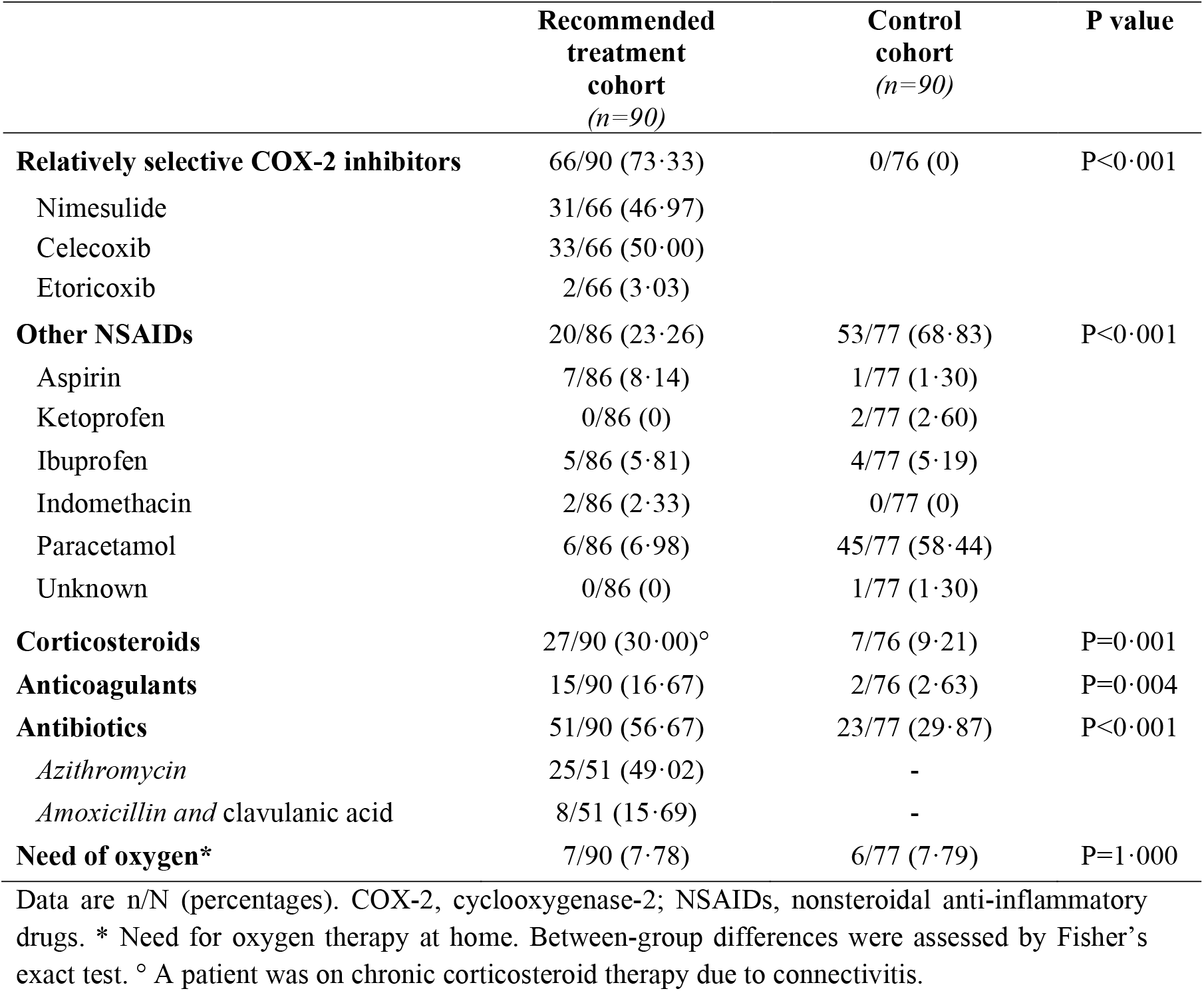
Treatment at home in the two study cohorts.

A sensitivity analysis of hospital admissions was repeated after excluding patients who spontaneously started treatment with paracetamol before contacting their family doctors in the ‘recommended’ cohort and the related matched patients in the ‘control’ cohort. Similarly to the intention-to-treat analysis, the event rate was still significantly lower in the ‘recommended’ than in the ‘control’ cohort (Log-rank test, p=0·0035) (Figure 2B).

In the ‘recommended’ cohort, anti-inflammatory treatment with NSAIDs started at home within a median of 2 days [IQR: 1-3] after the onset of COVID-19 symptoms. In both cohorts, all patients achieved complete remission, defined as resolution of major symptoms (Table 4). Nonetheless, symptoms like anosmia, ageusia/dysgeusia, lack of appetite and fatigue persisted in a lower percentage of patients in the ‘recommended’ than in the ‘control’ cohort (23·3% vs 73·3%, respectively, p<0·0001). In particular, this significant difference was documented in the subgroups of patients in whom these symptoms persisted for less than 30 days or more than 60 days (Table 4).

**Table 4.** Secondary outcomes.

|  | **Recommended treatment cohort**<br>**(n=90)** | **Control cohort**<br>**(n=90)** | **P value nominal** |
| --- | --- | --- | --- |
| Time from symptoms onset and start of anti-inflammatory therapy (days) | 2 [1-3] | - | - |
| Rate of complete remission* | 90/90 (100) | 90/90 (100) | P=1·000 |
| Rate of partial remission<sup>°</sup> | 21/90 (23·3) | 66/90 (73·3) | P<0·0001 ** |
| Persistence of minor symptoms (days) |  |  |  |
| < 30 | 11/21 (52·4) | 13/65 (20·0) | P=0·0107 |
| 30-60 | 5/21 (23·8) | 16/65 (24·6) |  |
| > 60 | 5/21 (23·8) | 36/65 (55·4) |  |
| Rate of hospitalisation | 2/90 (2·2) | 13/90 (14·4) | P=0·0053 ** |
| Rate of hospitalisation<sup>§</sup> | 1/84 (1·2) | 11/84 (13·1) | P=0·007 ** |

Data are n/N (percentages) or median [interquartile range], as appropriate. \* defined as complete recovery from major symptoms, ie no fever, SpO<sub>2</sub> >94% and/or no dyspnea, no cough, no rhinitis, no pain (myalgia, arthralgia, chest pain, headache, sore throat), no vertigo, no nausea, vomiting or diarrhoea, no sicca syndrome or red eyes; <sup>°</sup> defined as recovery from major COVID-19 symptoms, but persistence of symptoms such as anosmia, ageusia/dysgeusia, lack of appetite, fatigue. <sup>§</sup> Sensitivity analysis performed excluding patients who spontaneously started treatment with paracetamol before contacting their family doctors in the “recommended” cohort and the related matched patients in the “control” cohort. \*\* Significant after Bonferroni adjustment for multiple tests.

## Discussion

In this fully academic observational, matched-cohort study we found that early treatment of COVID-19 patients at home by their family doctors according to the proposed recommendation regimen almost completely prevented the need for hospital admission (the most clinically relevant outcome) due to progression toward more severe illness, compared to patients in the ‘control’ cohort who were treated at home according to their family physician’s assessments. This translated into a reduction of over 90% in the overall numbers of days of hospitalisation and in related treatment costs. Considering that differences in early at-home treatment regimens were negligible, the cost effectiveness of the home therapy algorithm was terrific. This was consistent with the finding that only 1.2 patients needed to be treated to prevent one hospitalisation event. Although the study failed to detect a significant treatment effect on time to complete remission of symptoms, the primary outcome of the study, it is noteworthy that the ‘recommended’ cohort required a few more days to reach the resolution of major early symptoms, including fever, musculoskeletal pain, headache, and cough, than in the ‘control’ cohort. Symptoms, such as anosmia or ageusia/dysgeusia, persisted less frequently and for a shorter period in the ‘recommended’ than in the ‘control’ cohort. Why treatment effect on risk of hospitalisation was so different from treatment effect on disease duration is a matter of speculation. One plausible explanation is that we were not testing disease-modifying treatments, but rather comparing different symptomatic regimens. In other words, the early home therapy regimen could not appreciably affect the duration of the diseases, but could affect disease phenotype, with a consequent, remarkably reduced need for hospitalisation. The results are even more surprising when one considers that controls presented with symptoms during the first wave of the epidemic, when the health care system was pushed to its limit and not all patients in need may have accessed the hospital because of severe limitations of available resources. Thus, the lower hospitalisation rate of patients given at-home therapy according to guidelines cannot be ascribed to limited access to hospitals.

The pillars of the proposed treatment recommendation ^10^ are three: i) intervene at the very onset of mild/moderate symptoms at home; ii) start treatment as early as possible after the family doctor has been called by the patient, without awaiting the results of a nasopharyngeal swab; iii) rely on specific non-steroidal anti-inflammatory drugs, unless contraindicated. Indeed, after the initial exposure to SARS-CoV-2, patients typically develop symptoms that indicate an inflammatory process within 5 to 6 days on average.^23^ Insights into the pathogenic mechanism underlying SARS-CoV-2 infection highlight the critical role of inflammatory hyper-response, characterised by tissue leucocyte infiltration, macrophage activation, widespread endothelial damage, complement–induced blood clotting and systemic microangiopathy, in disease progression.^24^ There is growing evidence to suggest that this hyper-inflammatory reaction, rather than the virus itself, underpins the progression to severe COVID-19 cases, and pro-inflammatory cytokines and macrophages seem to be integral to the initiation and propagation of this process.^24^ Therefore, the recommendation to start treating early COVID-19 symptoms with NSAIDs, whose best characterised mechanism of action is the inhibition of the cyclooxygenase (COX) activity of prostaglandin H synthase 1 and 2, also referred to as COX-1 and COX-2.^25^ COX-2 has a great effect on pro-inflammatory cytokines and its inhibition does not blunt immune response against viral disease.^11^ The COX-2 selectivity of a particular drug is a continuous variable in relation to the relative drug concentration required to inhibit COX-1 and COX-2 enzymes in whole blood assays by 50%.^25^ Substantial overlap in COX-2 selectivity is found among some coxibs (e.g., celecoxib) and some traditional NSAIDs (e.g., nimesulide).^25^ The experimental evidence that celecoxib decreased cytokine levels (TNF-α, G-CSF and IL-6) in bronchoalveolar lavage fluid in mice with influenza A infection,^26^ and the overlap in COX-2 selectivity between this coxib and nimesulide, was the rationale for recommending these two drugs for the treatment of early COVID-19 symptoms at home, if not contraindicated.

Adherence to this recommendation was high (73·3%) in the ‘recommended’ cohort. Conversely, we found that in the ‘control’ cohort, none of the patients received a COX-2 inhibitor, and most were given paracetamol, a drug with very mild anti-inflammatory activity.^27^ Paracetamol is suggested as a safe and recommendable alternative for the early management of pain and fever in COVID-19 patients. However, it should be taken into account that besides being a negligible anti-inflammatory drug, paracetamol reduces plasma and tissue gluthatione levels when given at relatively low doses, which might exacerbate COVID-19, as recently hypothesised.^28^ Although more selective inhibition of COX-2 is desirable to limit the gastrointestinal toxicity seen with less selective COX-2 inhibitors, physicians may be aware of the finding that the use of NSAIDs has been associated with higher rates of cardiovascular events.^29^ Moreover, nimesulide can be associated with a risk of hepatotoxicity, which is very low when the drug is administered at the recommended time and daily dosage.^30^ Nonetheless, in the ‘recommended’ cohort, treatment with nimesulide or celecoxib was safe and well tolerated, with only one patient reporting epigastric pain. This may explain the low rate of the use of aspirin in this cohort, which according to the proposed recommendations should be given as an alternative treatment to nimesulide and celecoxib when signs of toxicity or contraindications to these drugs are brought to the attention of the family physician. Nonetheless, aspirin could be a potential alternative treatment for COVID-19 at home, since it has been shown to reduce plasma levels of inflammatory cytokines in patients with chronic stable angina,^31^ and even to have antiviral activity against RNA viruses of the respiratory tract.^32^ The treatment effect of this drug is supported by the findings of a retrospective cohort study on 412 adult patients hospitalised with COVID-19, which showed that aspirin administration was independently associated with a reduced risk of mechanical ventilation, intensive care unit admission, and in-hospital mortality.^33^

According to the recommendation algorithm, corticosteroids were not used at the onset of symptoms but only after a mean of 8 days in 30% of patients in the ‘recommended’ cohort in whom fever, myalgia/arthralgia or cough persisted. A patient in this cohort was already receiving corticosteroids chronically due to connectivitis. Indeed, corticosteroids exert their anti-inflammatory effects mainly by inhibiting pro-inflammatory genes that encode for cytokines, chemokines, inflammatory enzymes to control the inflammatory process and restore homeostasis.^34^ However, the use of corticosteroids in COVID-19 patients has been controversial, due to the risk of prolonging the presence of the virus in the respiratory tract and blood, and the incidence of complications, as shown in previous observational studies in patients with coronavirus pneumonia induced by SARS and MERS.^35,36^ Nevertheless, none of the patients in the ‘recommended’ cohort given corticosteroids exhibited any particular side effects related to the use of these medicines. Based mainly on the positive findings of reduced mortality in hospitalised patients in the large RECOVERY trial, WHO guidance strongly recommended systemic corticosteroids in patients with severe COVID-19, except in those who were not receiving respiratory support, who did not benefit from the treatment.^37^ Data for the early phase of COVID-19, when patients are not hospitalised, are scanty, but some evidence indicates that prompt intervention with corticosteroids can reverse or at least attenuate the initial lesions in the lungs.^12,38^

Apart from causing patients to be bedridden even with mild symptoms, there is evidence that in SARS-CoV-2 infection, dysregulation of the coagulation cascade and fibrinolytic systems occur, creating a high risk of thromboembolic events and death for patients.^39^ Thus, the use of low-molecular weight (LMW) heparin at a prophylactic dose has been recommended for the management of COVID-19 patients. However, only 16% of patients in the ‘recommended’ cohort were treated prophylactically with LMW heparin because bedridden, without side effects. This suggests the need for further educational programmes for family physicians on this topic.

The use of antibiotics in non-hospitalised COVID-19 patients is not mandatory, but sometimes necessary, since there is evidence that patients may die of secondary bacterial infections rather than viral infection. Thus, as indicated in the proposed recommendations, antibiotics were prescribed to patients in both cohorts only when needed, not on a routine basis. This is in line with the very recent findings of the PRINCIPLE trial, which do not justify the routine use of azithromycin for shortening time to recovery or reducing the risk of hospitalisation in individuals with suspected COVID-19 illness in the community.^40^ This has important implications, since indiscriminate use of antibiotics could favour the development of antimicrobial resistance.

### Limitations and strengths

We failed to demonstrate any treatment effect on time to resolution of symptoms (time to complete remission) ^21^ that was the primary outcome of the study. The relatively small sample size was not an explanation of this negative finding because time to resolution of major COVID-19 symptoms observed in our controls was consistent with the assumptions used for power calculation. In actual facts, the time to complete remission of symptoms in the two cohorts was quite similar. This finding could be explained by the fact that the tested treatments were targeting symptoms and were not specific to the virus. Therefore, it could be speculated that the time of viral clearance would be comparable in the two cohorts, independently of the symptomatic therapy used, but symptoms would be attenuated to the extent of not requiring hospital admission. Other major limitations included the non-randomised design and the retrospective nature of statistical analyses. However, study analyses were performed according to the predefined study protocol and statistical plans. At variance with data in the ‘recommended’ cohort collected by family physicians, the outcome data of the ‘control’ cohort were obtained from patient questionnaires and interviews referring to events that had occurred many months before the survey, which may have resulted in an underestimation of time to resolution of COVID-19 symptoms and of adverse event rates, but not on the hospitalisation rate. Indeed, the date of hospital admission was well documented by the hospital discharge letter.

Moreover, data from the ‘control’ cohort were obtained when hospitals were under huge pressure because of the first ‘wave’ of the COVID-19 pandemic, which may have resulted in postponed or avoided hospitalisation of patients in need. Findings of remarkably higher hospitalisation rates in the ‘control’ cohort of patients, despite this potential bias, provided additional, indirect evidence, of the protective effect of the proposed recommended treatment protocol against hospitalisation because of worsening of COVID-19 symptoms. However, time to hospitalization was a secondary outcome of the study and the sample size was not calculated on the basis of an expected treatment effect on this outcome. Thus, the possibility of a casual finding cannot be definitely excluded and the observed reduction in patients hospitalizations should be considered as an hypothesis generating finding that could provide a robust background for a prospective trial primarily aimed to test treatment effect on this outcome.

The proposed recommendation algorithm suggests upgrading treatment toward the use of corticosteroids or to start anticoagulant prophylaxis, based also on hematochemical tests that document any increases in inflammatory indexes (CRP, neutrophil count) and/or D-dimer, respectively, in addition to clinical judgement. However, fulfilling this lab test requirement in the early phase of the illness was not feasible, since all patients had confirmation of SARS-CoV-2 infection and were thus quarantined at home, making it impossible for them to reach the laboratory. The strengths of the COVER study include the formal evaluation of a treatment recommendation algorithm for family doctors targeting early symptoms in the community, designed according to a pathophysiologic and pharmacologic rationale. Several recommendations on how to treat COVID-19 patients at home have recently been proposed, including those of the Italian Ministry of Health,^41^ but none have been formally tested for their ability to prevent or limit the progression of the early phase of the illness to the need for hospitalisation.

In conclusion, we found that a few simple treatments, as reported in the proposed recommendation algorithm, show benefits among outpatients in the early phase of COVID-19. This reasoned approach may have the potential to avert clinical deterioration of the illness, limiting the need for hospitalisation, in addition to shortening the duration of symptoms, such as anosmia, dysgeusia and fatigue, which affect patients’ quality of life, with substantial public health and societal implications and effects. Results of these retrospective analyses could provide the background and a hypothesis-generating finding for designing future prospective trials in this context.

## Data Availability

Sharing of individual participant data with third parties was not specifically included in the informed consent of the study, and unrestricted diffusion of such data may pose a potential threat of revealing participants identities, as permanent data anonymization was not carried out (patient records were instead de-identified per protocol during the data retention process). To minimize this risk, individual participant data that underlie the results reported in this article will be available after three months and up to five years from article publication. Researchers shall submit a methodologically sound proposal to Dr. Annalisa Perna, head of the Laboratory of Biostatistics of the Department of Renal Medicine of the Istituto di Ricerche Farmacologiche Mario Negri IRCCS. To gain access, data requestors will need to sign a data access agreement and obtain the approval of the local ethics committee.

## Contributing Authors

FS and GR had the original idea; NP and GR wrote the original version of the manuscript; EC, SP, CM, EP, MVP, GP, UC, FS contributed to patient identification; NR helped in data collection and managing; AP, TP performed the statistical analyses; NP, PR, GR elaborated the final version of the manuscript, all authors critically revised the final version. NP and GR took the responsibility for the submission for publication. No medical writer was involved.

## Data Sharing

Sharing of individual participant data with third parties was not specifically included in the informed consent of the study, and unrestricted diffusion of such data may pose a potential threat of revealing participants’ identities, as permanent data anonymisation was not carried out (patient records were instead de-identified per protocol during the data retention process). To minimize this risk, individual participant data that underlie the results reported in this article will be available after three months and up to five years from article publication. Researchers shall submit a methodologically sound proposal to Dr. Annalisa Perna, head of the Laboratory of Biostatistics of the Department of Renal Medicine of the Istituto di Ricerche Farmacologiche Mario Negri IRCCS. To gain access, data requestors will need to sign a data access agreement and obtain the approval of the local ethics committee.

## SUPPLEMENTARY METHODS

### Summary of proposed recommendations for treatment of COVID-19 patients at home

Recommended treatments should start immediately when COVID-19 early symptoms appear without waiting results of a nasopharyngeal swab, if any. The recommended drugs can be used unless contraindicated according to the summary of product characteristics.

I. <u>Non-steroidal anti-inflammatory drugs (NSAIDs)</u> *<u>Relatively selective COX-2 inhibitors</u>* ^§#^ (*for myalgias and/or arthralgias or other painful symptoms*) ^*§*^ *based on the ratio of concentrations of the various NSAIDs required to inhibit the activity of COX-1 and COX-2 by 50 percent (IC*_*50*_*) in assays of whole blood* *#unless contraindicated Nimesulide \** 100 mg b.i.d p.o, after a meal, for a maximum of 12 days. *Or Celecoxib \** Initial oral dose of 400 mg, followed by a second dose of 200 mg on the first day of therapy. In the following days, up to a maximum of 400 mg (200 mg twice a day) should be given as needed for a maximum of 12 days *\* Should the patient have fever (*≥ *37*.*3 °C) or develop laboratory signs of hepatotoxicity associated with nimesulide or there are contraindications to celecoxib, these drugs should be substituted with aspirin (a COX-1 and COX-2 inhibitor) (500 mg twice a day p*.*o. - after a meal). These treatments should be associated with a proton pump inhibitor (e*.*g. lansoprazole - 30 mg/day; or omeprazole - 20 mg/day; or pantoprazole - 20 mg/day)*. *After approximately 3 days from the onset of symptoms (or more days are elapsed and the physician sees the patient for the first time), a series of hematochemical tests should be performed (blood cell count, D-dimer, CRP, creatinine, fasting blood glucose, ALT). Should inflammatory indexes (CRP, neutrophil count), ALT, and D-dimer be in the normal range, nimesulide****/****celecoxib (or aspirin) treatment will continue*.
II. <u>Corticosteroids*</u> <u>Dexamethasone</u> (*for persistent fever or musculoskeletal pain or when few days later hematochemical tests were repeated and even mild increase of inflammatory indexes - CRP, neutrophil count - are documented, or cough and oxygen saturation (SpO*_*2*_*) <94-92% occur)* 8 mg p.o for 3 days, then tapered to 4 mg for a further 3 days, and then to 2 mg for 3 days. That makes 42 mg dexamethasone total over 9 days. \**Duration of corticosteroid treatment also depends on the clinical evolution of the disease*
III. <u>Anticoagulants</u> *<u>Low-molecular weight (LMW) heparin</u>*\* *(when the hematochemical tests show even a mild increase of D-dimer or for thromboembolism prophylaxis for bedridden patients)* Enoxaparin, at the prophylactic daily dose of 4000 U.I subcutaneously - i.e. 40 mg enoxaparin. Treatment recommended for at least 7-14 days, independently of the patient recovering mobility. \**unless contraindicated (e*.*g*., *ongoing bleeding or platelet count <25 x 10*^*9*^*/L)*
IV. <u>Oxygen therapy</u> Gentle oxygen supply in the early phase of the disease, possibly before pulmonary symptoms manifest, in the presence of progressively decreasing oxygen saturation – as indicated by oximeter – or following a first episode of dyspnoea or wheezing. Conventional oxygen therapy is suggested when the respiratory rate is >14/min and oxygen saturation (SpO_2_) *<94-92%*, but is required with SpO_2_ <90% at room air. With liquid oxygen, start with 8-10 litre/min and monitor SpO_2_ every 3-4 hours. Titrate oxygen flow rate to reach target SpO_2_ *>94%*. Then the rate of oxygen administration can be reduced to 4-5 litre/min (but continue SpO_2_ monitoring every 3-4 hours). With gaseous O_2,_ start with 2.5-3.0 litre/min, but monitor SpO_2_ more frequently than with liquid oxygen, and titrate flow rates to reach target SpO_2_ *>94%*. Should patients be poorly responsive to high O_2_ administration, consider hospitalisation, if feasible.
V. <u>Antibiotics</u> *<u>Azithromycin</u>*\* *(with bacterial pneumonia or suspected secondary bacterial upper respiratory tract infections, or in particularly fragile patients, or when hematochemical inflammatory indexes (CRP, neutrophil count) are markedly altered)* 500 mg/day p.o for 6-10 days depending on the clinical judgement * *Should the patient be at risk of or with a history of cardiac arrhythmia or present other contraindications, cefixime (400 mg/day p*.*o for 6-10 days) or amoxicillin/clavulanic acid (1gr three times a day for 6-10 days) can be considered as alternative to azithromycin*.

### COVER Study Organization

Members of the COVER Study Organization were as follows (all in Italy): *Chief Investigator* - Giuseppe Remuzzi (Bergamo); *Study coordinators* - Norberto Perico, Fredy Suter (Bergamo); *Coordinating Centre* – Istituto di Ricerche Farmacologiche Mario Negri IRCCS, Centro di Ricerche Cliniche per le Malattie Rare *Aldo e Cele Daccò*, Ranica (Bergamo); *Study investigators including patients* - Elena Consolaro (Varese), Chiara Moroni (Varese), Umberto Cantarelli (Teramo), Stefania Pedroni (Varese), Maria Vittoria Paganini (Varese), Elena Pastò (Varese), Grazia Pravettoni (Varese), Fredy Suter (Bergamo); *Data collection and processing* - Nadia Rubis, Davide Villa, Olimpia Diadei, Sergio Carminati (Bergamo); *Data Analysis* - Annalisa Perna, Tobia Peracchi (Bergamo); *Regulatory Affairs* - Paola Boccardo (Bergamo); *Finalization of the manuscript*: Norberto Perico, Piero Ruggenenti, Giuseppe Remuzzi (Bergamo).

